# Detectability of the novel coronavirus (SARS-CoV-2) infection and rates of mortality for COVID-19 in different regions of the Russian Federation

**DOI:** 10.1101/2020.09.18.20197194

**Authors:** Edward Goldstein

## Abstract

**Relevance:** Laboratory diagnosis of the novel coronavirus (SARS-CoV-2) infection combined with tracing/quarantine for contacts of infected individuals affects the spread of the SARS-CoV-2 infection and levels of related mortality. At the same time, testing practices for SARS-CoV-2 infection vary in the different regions of the Russian Federation. For example, in the city of St. Petersburg, where mortality rate for COVID-19 is the highest in the Russian Federation on Oct. 25, 2020, every death for COVID-19 corresponds to 15.7 detected cases of COVID-19 in the population, while the corresponding number for the whole of Russia is 58.1, suggesting limited detection of mild and moderate cases of COVID-19 in St. Petersburg. Additionally, while in some regions in Russia, all individuals with respiratory symptoms presenting for medical care are tested for SARS-CoV-2 infection, in a number of other regions in Russia, only certain categories of individuals presenting for medical care with respiratory symptoms were tested for the SARS-CoV-2 infection prior to Oct. 26, 2020.

**Materials & Methods:** More active testing for SARS-CoV-2 in the population results in increased detectability (i.e. the proportion of detected COVID-19 cases among all cases of SARS-CoV-2 infection in the population) and decreased case-fatality ratio (CFR, the proportion of deaths among reported COVID-19 cases in the population) – this because under more active testing, the number of mild and moderate cases of COVID-19 increases. We used data from the Russian Federal Service for Surveillance on Consumer Rights Protection and Human Wellbeing (Rospotrebnadzor) on the number of detected cases and the number of deaths from COVID-19 in the different regions of the Russian Federation to examine the correlation between case-fatality ratios and rates of mortality for COVID-19 in different regions of the Russian Federation.

**Results:** The correlation between case-fatality ratios and rates of mortality for COVID-19 in the 85 different regions of the Russian Federation on Oct. 25, 2020 is 0.64 (0.50,0.75). For several regions of the Russian Federation, detectability of SARS-CoV-2 infection is relatively low, while rates of mortality for COVID-19 are relatively high.

**Conclusions:** Detectability of the SARS-CoV-2 infection is one of the factors that affects the levels of mortality from COVID-19 – higher detectability contributes to lower rates of mortality for COVID-19. To increase detectability, one ought to test all individuals with respiratory symptoms seeking medical care for SARS-CoV-2 infection (which is also suggested by the recent recommendations from the Ministry of Health), and to undertake additional measures to increase the volume of testing for SARS-CoV-2. Such measures, in combination with quarantine for infected cases and their close contacts help to mitigate the spread of the SARS-CoV-2 infection and diminish the related mortality.

## Introduction

Laboratory diagnosis of the novel coronavirus (SARS-CoV-2) infection combined with tracing/quarantine for contacts of infected individuals is an effective means for diminishing the spread of SARS-CoV-2 in the community and decreasing the levels of associated mortality. For example, the corresponding diagnostics and quarantine is widely practiced in Iceland -- using serological data and data on PCR-detected SARS-CoV-2 infections, Gudbjartsson et al. estimated that 56% of all cases of SARS-CoV-2 infection in Iceland were PCR-detected [1]. During the Spring wave of the SARS-CoV-2 epidemic in New South Wales, Australia, a small number of infections were recorded in schools [2], with widespread testing in schools and subsequent quarantine for contacts of detected cases practiced. Widespread testing for SARS-CoV-2 infection is also conducted in the Russian Federation [3]; however, testing practices for SARS-CoV-2 infection vary in the different regions of the Russian Federation. For example, while in some regions in Russia, all individuals with respiratory symptoms presenting for medical care are tested for SARS-CoV-2 infection [4,5], in a number of other regions in Russia, only certain categories of individuals presenting for medical care with respiratory symptoms (e.g. persons aged over 65y, healthcare workers, etc.) were tested for the SARS-CoV-2 infection prior to Oct. 26, 2020 [6,7]. There is limited information on the effect of differences in testing practices on the spread of SARS-CoV-2 infection and the rates of related mortality.

The effect of testing practices on the spread of SARS-CoV-2 infection depends on the detectability of the SARS-CoV-2 infection (i.e. the proportion of detected COVID-19 cases among all cases of SARS-CoV-2 infection in the population). Higher detectability in combination with quarantine for infected individuals and their (symptomatic) contacts results in the prevention of a greater number of new cases of infection, bringing down the rate of spread of the SARS-CoV-2 infection in the population. Under more active testing for SARS-CoV-2 in the population, detectability of SARS-CoV-2 infection increases, and the case-fatality ratio (i.e. the proportion of deaths among reported COVID-19 cases in the population) decreases – this because under more active testing, the number of detected mild and moderate cases of COVID-19 increases, with the ratio between the number of deaths and the number of cases declining. Therefore, a lower case-fatality ratio generally corresponds to higher detectability of SARS-CoV-2 infection in the different regions in Russia. Here, we used data from the Russian Federal Service for Surveillance on Consumer Rights Protection and Human Wellbeing (Rospotrebnadzor) on the number of detected cases of COVID-19 and the number of deaths from COVID-19 in the different regions of the Russian Federation [8] to examine the relation between case-fatality ratios and rates of mortality for COVID-19 in the 85 different regions of the Russian Federation (Federal subject of Russia).

## Methods

### Data

We used data from Rospotrebnadrzor on the number of detected COVID-19 cases and the number of deaths from COVID-19 reported by Oct. 25, 2020 in the 85 different regions of the Russian Federation [8], as well as data from the Russian Federal State Statistics Service (Rosstat) on the population on January 1, 2020 for the 85 regions of the Russian Federation [9].

#### Informed consent

We used publicly available, aggregate data, with no informed consent from the participants sought.

### Statistical analysis

We evaluated the linear correlation between case-fatality ratios for COVID-19 for cases/deaths reported by Oct. 25, 2020 and rates of mortality for COVID-19 per 100,000 persons for deaths reported by Oct. 25, 2020 in the 85 different regions of the Russian Federation.

## Results

Figure 1 shows the case-fatality ratios (i.e. the proportion of deaths among reported COVID-19 cases in the population by Oct. 25, 2020) and rates of mortality for COVID-19 per 100,000 persons for deaths reported by Oct. 25, 2020 in the 85 regions of the Russian Federation. The correlation between case-fatality ratios and rates of mortality for COVID-19 in the different regions of the Russian Federation on Oct. 25, 2020 is 0.64 (0.50,0.75). The region with both the highest COVID-19 mortality rate per 100,000 persons and the highest case-fatality ratio – 6.4% (lowest detectability of SARS-COV-2 infection) is the city of St. Petersburg (top right of Figure 1). For each COVID-19 death in St. Petersburg reported by Oct. 25, 2020, there are 15.7 detected COVID-19 cases, compared to a national average of 58.1 cases, suggesting limited detectability of SARS-CoV-2 infection in St. Petersburg.

**Figure 1:**
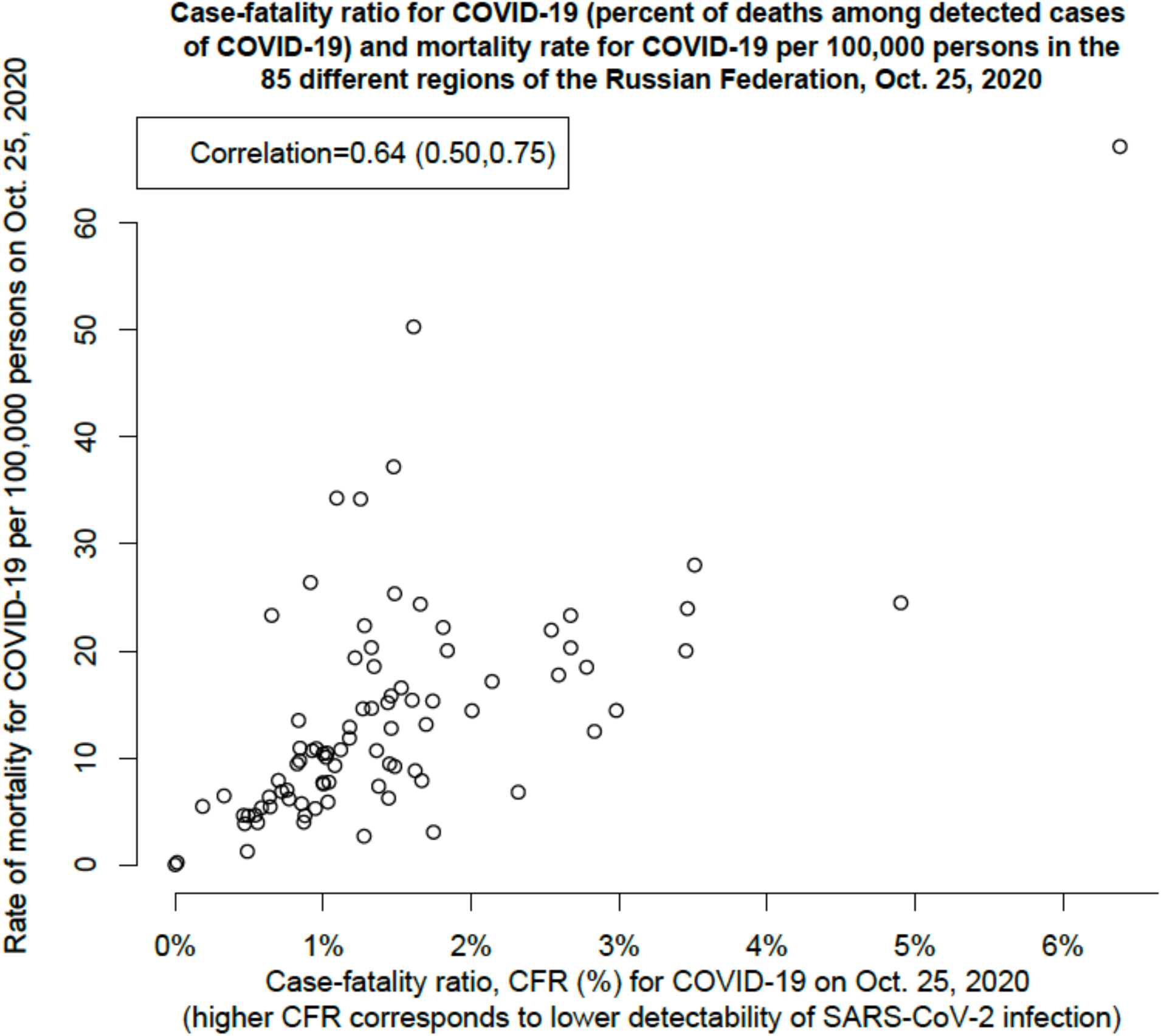
Case-fatality ratios (percent of deaths among reported COVID-19 cases in the population by Oct. 25, 2020) and rates of mortality for COVID-19 per 100,000 persons for deaths reported by Oct. 25, 2020 in the 85 regions of the Russian Federation.

## Discussion

Active testing for the novel coronavirus (SARS-CoV-2) infection combined with tracing/quarantine for close contacts of infected individuals affects the spread of SARS-CoV-2 [10] and the levels of related mortality. High levels of testing for the novel coronavirus infection were reported in several countries [1,11], including the Russian Federation. Such robust testing for SARS-CoV-2 leads to high detectability of SARS-CoV-2 infection (i.e. the percent of detected COVID-19 cases among all cases of SARS-CoV-2 infection in the population) – for example, in Iceland, detectability of SARS-CoV-2 infection was estimated at 56% [1]. Testing practices for SARS-CoV-2 infection vary in the different regions of the Russian Federation. For example, while in some regions in Russia, all individuals with respiratory symptoms presenting for medical care are tested for SARS-CoV-2 infection [4,5], in a number of other regions in Russia, only certain categories of individuals presenting for medical care with respiratory symptoms (e.g. persons aged over 65y, healthcare workers, etc.) were tested for the SARS-CoV-2 infection prior to Oct. 26, 2020 [6,7]. New guidelines, including the recommendation for testing of all symptomatic individuals for SARS-CoV-2 infection were issued by the Russian Ministry of Health on Oct. 26, 2020 [3], and the effect of those guidelines on testing practices in still uncertain. Moreover, there is limited information on the effect of (prior) differences in testing practices and detectability of SARS-CoV-2 infection on the spread of SARS-CoV-2 and the rates of mortality for COVID-19 for the different regions of the Russian Federation.

Here, using data from Rospotrebnadrzor on the number of detected COVID-19 cases and the number of deaths for COVID-19 [8] we established a high correlation between case-fatality ratios (the proportion of deaths among reported COVID-19 cases in the population) and rates of mortality for COVID-19 in different regions of the Russian Federation. We should note that a lower case-fatality ratio corresponds to a higher detectability of SARS-COV-2 infection.

Correspondingly, testing for SARS-CoV-2 infection and detectability of SARS-CoV-2 infection are some of the factors that affect the levels of mortality for COVID-19 in Russia. In particular, higher detectability of SARS-CoV-2 infection is generally associated with lower mortality for COVID-19 (Figure 1). To increase detectability, one ought to test all individuals with respiratory symptoms seeking medical care for SARS-CoV-2 infection (which is also suggested by the recent recommendations from the Ministry of Health [3]), and to undertake additional measures to increase the volume of testing for SARS-CoV-2. Such measures, in combination with quarantine for infected cases and their close contacts help to mitigate the spread of the SARS-CoV-2 infection and diminish the related mortality.

## Data Availability

Data used in this analysis are publicly available thorugh the following URLs in ref. 3,4: 
https://yandex.ru/maps/covid19?ll=87.127143%2C49.616265&z=3
https://showdata.gks.ru/report/278928/

https://yandex.ru/maps/covid19?ll=87.127143%2C49.616265&z=3

https://showdata.gks.ru/report/278928/

